# Prevalence of diabetic macular edema and risk factors among diabetic patients at the University of Gondar Tertiary Eye Care and Training Center, North West Ethiopia

**DOI:** 10.1101/2022.04.13.22273816

**Authors:** Endale Kabtu, Asamere Tsegaw

## Abstract

**BACKGROUND:** Diabetic macular edema (DME) is the most common cause of visual impairment in patients with diabetes mellitus. The prevalence of DME globally is around 6.8 % and in Ethiopia range from 5.7% to 11%.Different factors are associated with DME including poor glycemic control, longer duration, hypertension, dyslipidemia

**OBJECTIVE:** To determine the prevalence and associated factors of diabetic macular edema among diabetic patients attending University of Gondar (UOG) hospital, tertiary eye care and training center, NW Ethiopia

**METHODS:** A hospital based cross-sectional study was conducted from March 2021 to October 2021. Socio-demographic, clinical and laboratory data of patients was gathered. The collected data was entered into epi-data 4.6 version, exported to SPSS version 20 and analyzed.

**RESULTS:** A total of 165 diabetic patients were enrolled with mean age of 54.71 ±13.66 years, 50.9% male, 85.5% urban dwellers, 79.9% type 2 DM, 49.7% on oral hypoglycemic agents and the mean duration of diabetes was 7.93 years. Cataract was the commonest ocular morbidity and 42% of patients had at least mild vision impairment. The overall prevalence of DME was 17% and 5.5% of patients had clinically significant macular edema (CSME). The presence of proteinuria was 8.04 times more likely to have DME.

**CONCLUSION:** The prevalence of DME among our patients was high. The presence of proteinuria was significantly associated with DME. Screening of diabetic patients for sight threatening retinopathy early and appropriate treatment is recommended.

## INTRODUCTION

Diabetic macular edema (DME) is the most common cause of visual reduction in patients with DM. It can occur in any stage of diabetic retinopathy. (1) Mechanism of DME is multi factorial and due to disruption of the blood--retinal barrier following hyperglycemia induced damage, which leads to increased accumulation of fluid within the retinal layers of the macula.(2)

The prevalence of DME globally is around 6.8 %.(1) In western societies the reported prevalence ranges from 3.8% to 11.1%. (1,3,4,5) The prevalence in Africa is reported to be higher and ranges from 8.0% Cameroon (6), 12.5% South Africa (7), 20.8% South Africa (8), 33.3% Kenya (9). In Ethiopia, the prevalence ranges from 5.7% to 11%.(10,11)

Different factors have been found to be associated with DME including type-I diabetes (12,13,14) poor glycemic control (15,16,17,18), longer duration of DM (1,3,12,16), systemic hypertension (3,8,13,16,17,18,19), dyslipidemia (12,20), insulin therapy (3,6), proteinuria (3,15,20) and cataract surgery (15,17,21).

There has not been any study that specifically evaluated risk factors to develop DME among our patients and the aim of our study was therefore to determine the prevalence and investigate risk factors that are associated with DME among diabetic patients attending in the study center.

## METHODS AND MATERIALS

### STUDY DESIGN AND PERIOD

A hospital based cross-sectional study was conducted at University of Gondar Tertiary eye care and training center from March 2021-October 2021.

### STUDY AREA

This study was conducted at University of Gondar tertiary eye care and training center which is a major ophthalmic center in Ethiopia. It is an ophthalmic referral center for the entire North-West Ethiopia of an estimated 14 million people. Over 50,000 patients are seen at the center annually as inpatient and outpatient basis. Currently there are 6 subspecialty clinics with 7 actively working ophthalmologists, 26 ophthalmology trainee residents, 38 optometrists, 35 general clinical nurses and ophthalmic nurses and other supporting staff working in the center.

#### Study Population

All diabetic patients who visited the tertiary eye care and training center during data collection period and fulfilled the inclusion criteria.

#### Inclusion Criteria

Medically diagnosed diabetic patients. Adequate visualization of the fundus is possible.

#### Exclusion criteria

Diabetic patients who had additional causes of macular edema Patients age below 18 years old.

### Data collection procedure

Semi-structured interviewer-administered questionnaire, document review, and ocular examination were used to collect data. The questionnaire consisted of three sections: sociodemographic variables (6 items), medical history (10 items), and checklist for clinical and laboratory data extraction (11 items). Data quality was ensured through pre-testing the questionnaire before the actual data collection period. Socio-demographic data and relevant medical history were filled into the pretested semi-structured questionnaire. Laboratory test results of a single record of the most recent fasting blood glucose (FBG) level, HgA1c, urine analysis, lipid profile were obtained. Blood pressure was measured in sitting position after 5–10 min of rest. Hypertension is defined as systolic BP of ≥140 mmHg and/or diastolic BP of ≥90 mmHg. (22) BMI was calculated from weight in kilograms and height in meters squared and categorized according to WHO classification. (23)

Best-corrected visual acuity was taken using Tumbling E Snellen visual acuity chart and patient sitting at 6 m position, and classified according to WHO grading of visual acuity (24) as follows: visual acuity better or equal to 6/18 – normal; visual acuity ≤6/24 and better than or equal to 6/60 – moderate visual impairment; visual acuity <6/60 and better than or equal to counting fingers at 3 m – severe visual impairment; visual acuity less than counting fingers at 3 m – blindness; the results for the eye with better visual acuity was recorded.

Anterior and posterior segment examinations were done using slit-lamp biomicroscope and 90D condensing lens was used for detailed evaluation of the retina after dilating the pupil with 1% tropicamide. Grading of the retinal changes was made using the Diabetic Retinopathy (DR) Study guidelines (25) and recorded in six categories: mild, moderate, and severe nonproliferative retinopathy and early, high risk, and advanced proliferative retinopathy. DME was diagnosed when there were hard exudates on the macula and/or macular thickening obvious on slit-lamp examination and clinically significant macular edema (CSME) was diagnosed based on ETDRS study criteria.(26) In cases of asymmetric involvement of eyes, the eye with the most severe DR grade was taken. In patients with concomitant central or branch retinal vein occlusion, the DR grade in the eye not involved in the vein occlusion was used. All data were collected and recorded by an ophthalmologist, and all diagnoses were confirmed by a retina specialist at the retina clinic of the study center.

### DATA PROCESSING AND ANALYSIS

The collected data was checked for accuracy and consistency and manual data clean up and correction of any errors was done. Data was coded and entered into epi-data4.6 and exported to statistical package for social sciences (SPSS) version 20 for analysis. Simple binary logistic regression analysis was done and the explanatory variables with pre-set p-value of <0.2 were taken for further analysis with multiple binary logistic regression to identify the factors independently associated with diabetic macula edema. Associations were shown in terms of calculated odds ratio and p-values. Results are described in terms of numbers, percentages, means and medians, and are displayed on tables, pie chart and bar graphs.

### ETHICAL CONSIDERATIONS

The study was conducted after ethical clearance was obtained from University of Gondar ethical review board (ID=UOG/ER/130/2022). Informed written consent was obtained from the study participants after clear explanation concerning the purpose and importance of the study. The identity of the patient was not exposed in any way and confidentiality of patient record was respected.

## RESULTS

A total of 165 diabetic patients were included in the study. The mean age was 54.71±13.66 years and range 19-87 years. A majority 84 (50.95%) were males and 141 (85.5%) were urban dwellers. (Table 1)

**Table-1.**
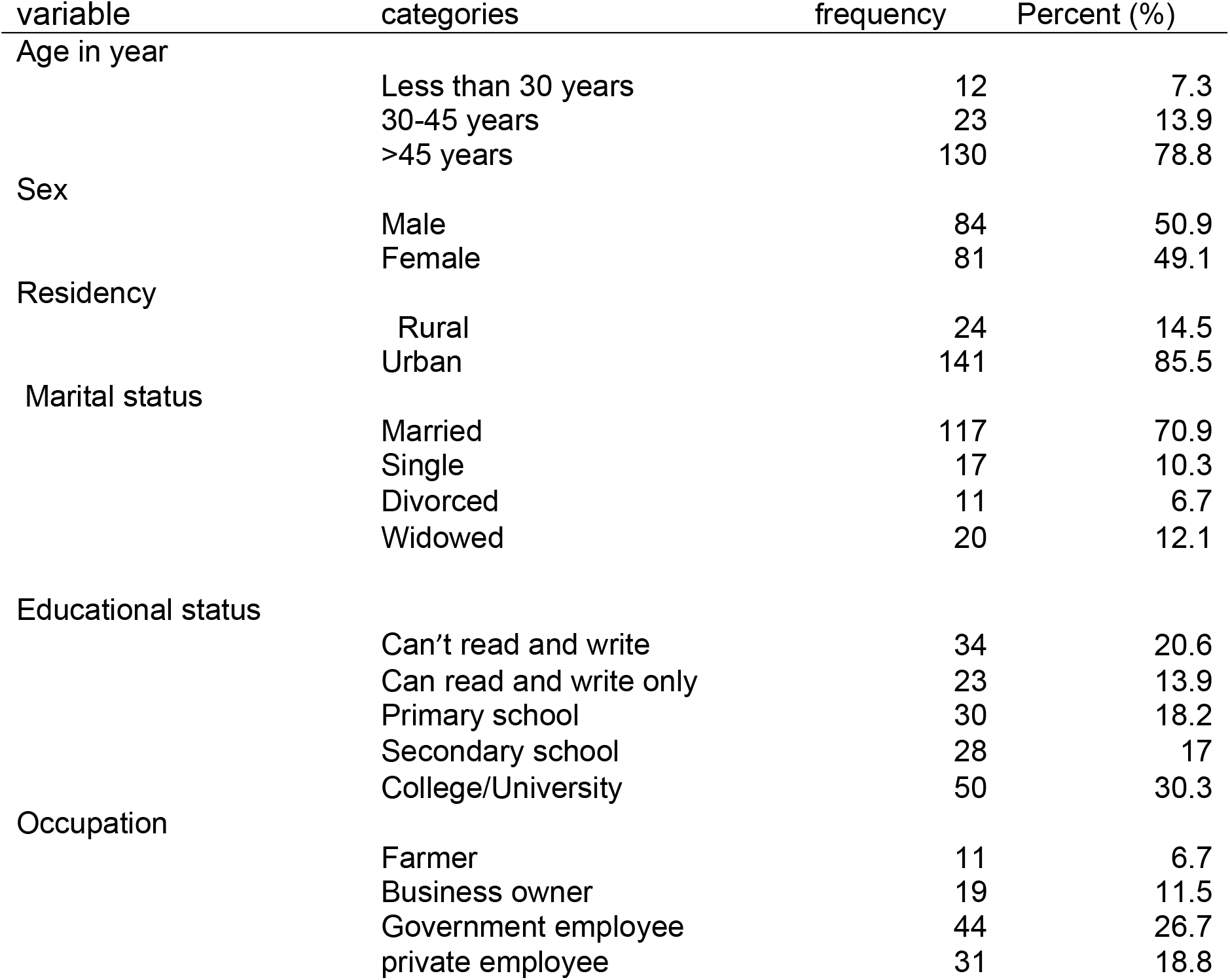
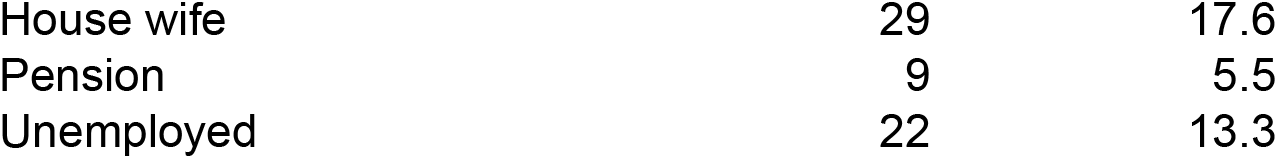
Socio-demographic characteristics of Diabetic patients presented to UOG tertiary eye care and training center, North West Ethiopia, 2021 (n=165)

| variable | categories | frequency | Percent (%) |
| --- | --- | --- | --- |
| Age in year | Less than 30 years | 12 | 7.3 |
|  | 30-45 years | 23 | 13.9 |
|  | >45 years | 130 | 78.8 |
| Sex | Male | 84 | 50.9 |
|  | Female | 81 | 49.1 |
| Residency | Rural | 24 | 14.5 |
|  | Urban | 141 | 85.5 |
| Marital status | Married | 117 | 70.9 |
|  | Single | 17 | 10.3 |
|  | Divorced | 11 | 6.7 |
|  | Widowed | 20 | 12.1 |
| Educational status | Can't read and write | 34 | 20.6 |
|  | Can read and write only | 23 | 13.9 |
|  | Primary school | 30 | 18.2 |
|  | Secondary school | 28 | 17 |
|  | College/University | 50 | 30.3 |
| Occupation | Farmer | 11 | 6.7 |
|  | Business owner | 19 | 11.5 |
|  | Government employee | 44 | 26.7 |
|  | private employee | 31 | 18.8 |

|  |  |  |
| --- | --- | --- |
| House wife | 29 | 17.6 |
| Pension | 9 | 5.5 |
| Unemployed | 22 | 13.3 |

Most of the patients had type-II DM 131 (79.4%), the mean duration of diabetes was 7.93 years (range 1-30 years) and a majority of them were on oral hypoglycemic agents 85 (49.7%). (Figure-I) (Table 2)

**Figure-I.**
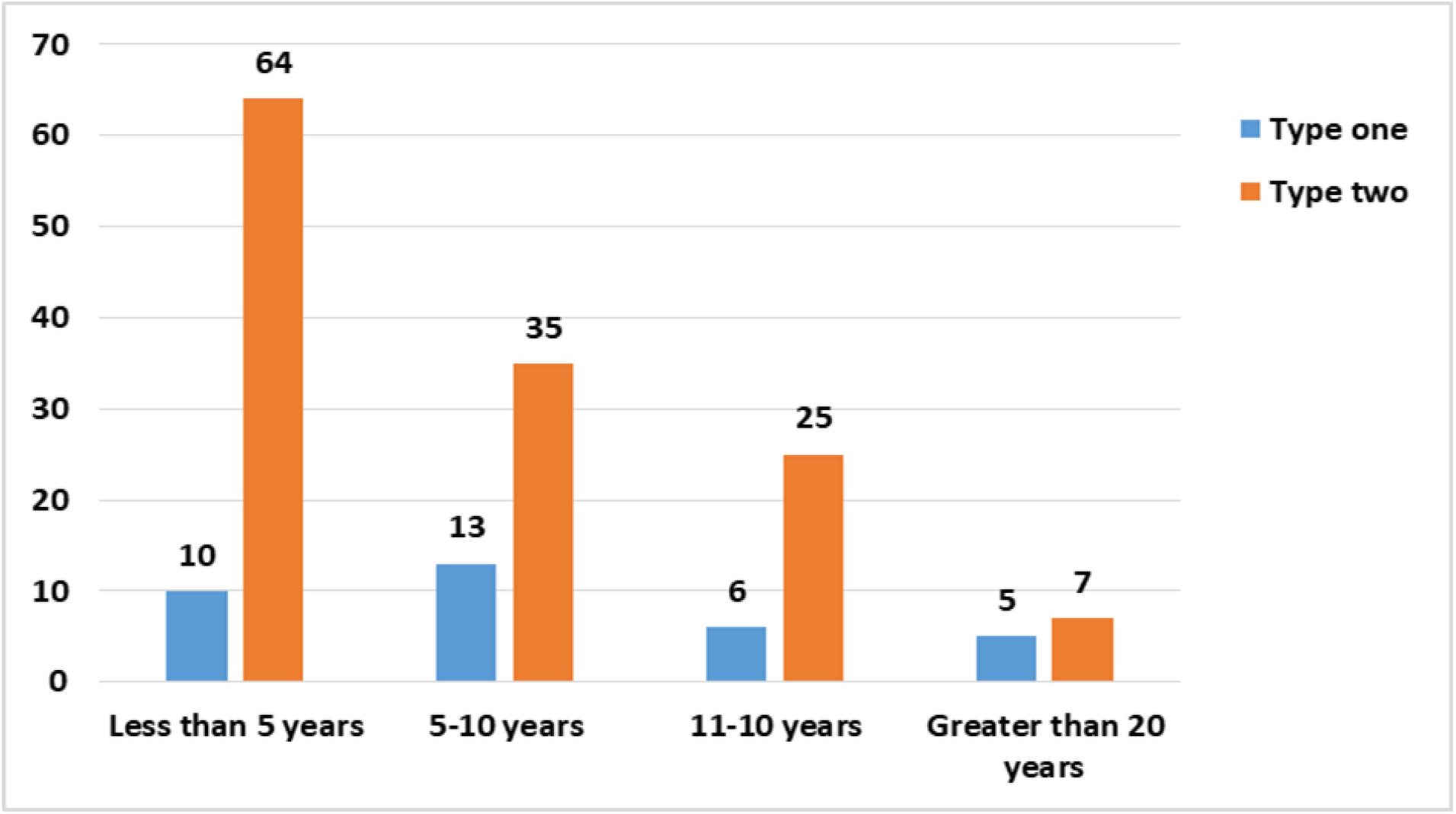
Disease duration among type I and type II diabetic patients at UOG tertiary eye care and training center, North West Ethiopia, 2021 (n=165)

**Table-II.** Clinical characteristics of diabetic patients presented to UoG tertiary eye care and training center, North West Ethiopia, 2021 (n=165)

| Variable | categories | Frequency<br>(number) | Percent (%) |
| --- | --- | --- | --- |
| Type of DM |  |  |  |
|  | Type 1 | 34 | 20.6 |
|  | Type 2 | 131 | 79.4 |
| Duration of DM |  |  |  |
|  | Less than 5 years | 74 | 44.8 |
|  | 5-10 years | 48 | 29.1 |
|  | 11-20 years | 31 | 18.8 |
|  | Greater than 20 years | 12 | 7.3 |
|  | Dietary | 10 | 6.1 |
| Form of DM therapy |  |  |  |
|  | OHA | 82 | 49.7 |
|  | Insulin | 50 | 30.3 |
|  | Combination | 23 | 13.9 |

Systemic hypertension was the most common known systemic co-morbidity 69 (41.8%) followed by dyslipidemia 22 (13.3%) and 21(12.7%) of patient had high systolic blood pressure measurements. (Table-III)

**Table-III.**
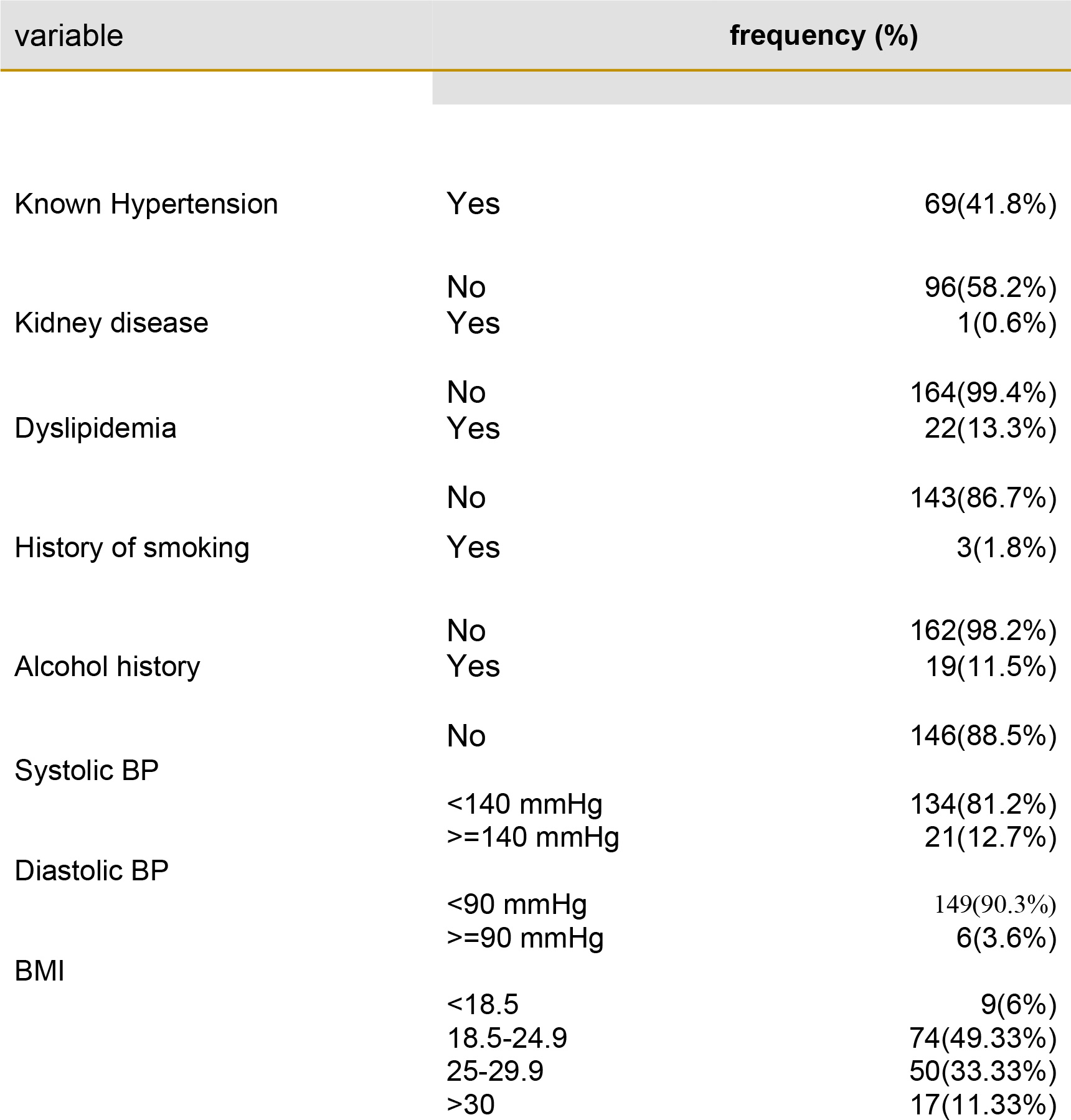
Concomitant systemic co-morbidities among patients among with diabetes at UOG tertiary eye care and training center, North West Ethiopia, 2021 (n=165)

Cataract was the most common concomitant ocular pathology 28 (17%), followed by Glaucoma 2(1.2%) and corneal opacity 2 (1.2%).

The means of FBS, HbA1c, cholesterol and triglycerides were 158.92 mg/dL, 8.86 mmol/mol, 178.96mg/dL and 175.01 mg/dl respectively. (Table-IV and Table-V)

**Table-IV.** Laboratory Investigation results (all in mg/dl except specified) of diabetic patients presented to UOG tertiary eye care and training center, North West Ethiopia, 2021 (n=65)

| Variable name | Minimum | maximum | Mean | SD |
| --- | --- | --- | --- | --- |
| FBS | 51 | 459 | 158.92 | ±68.36 |
| HbA1c<br>(mmol/mol) | 6.7 | 12.6 | 8.86 | ±1.38 |
| Total cholesterol | 56 | 345 | 178.96 | ±53.72 |
| Triglyceride | 46 | 490 | 175.01 | ±78.62 |
| LDL | 25 | 220 | 99.82 | ±35.06 |

**Table-V.**
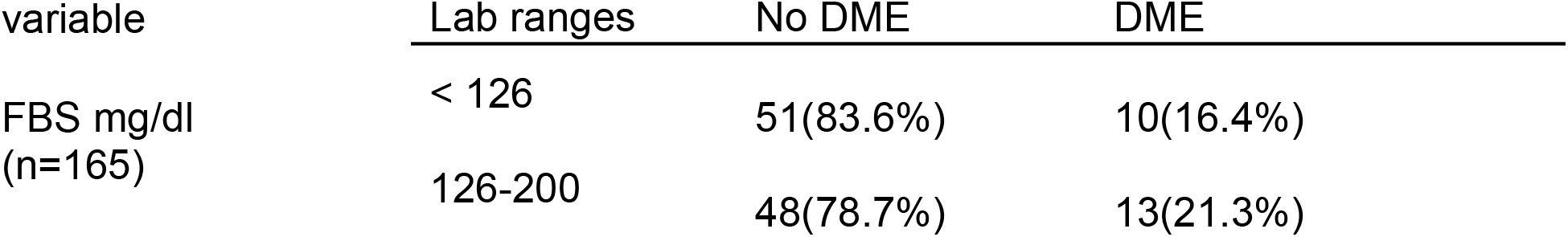
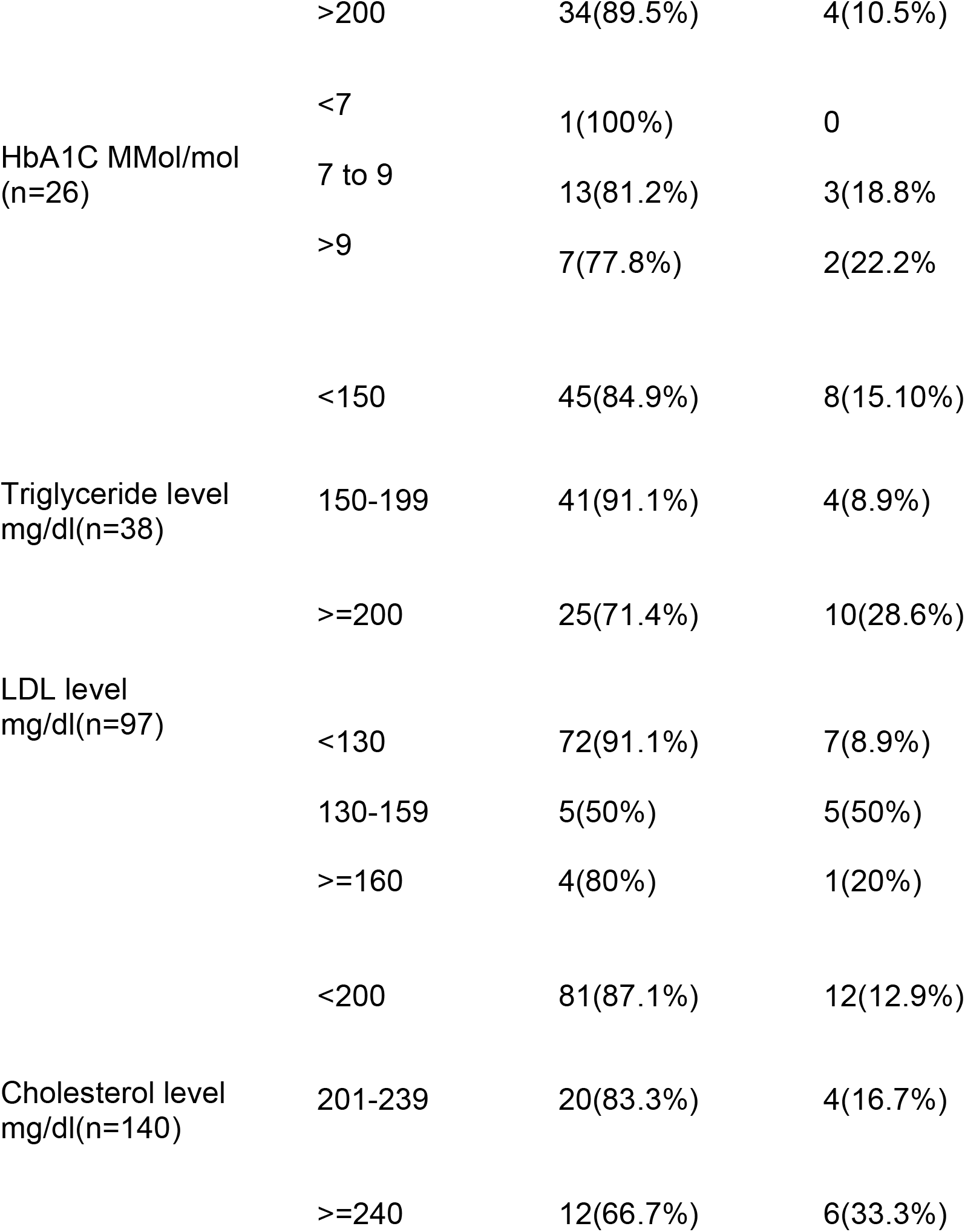
Category of Laboratory Investigation results versus the presence or absence of DME among diabetic patients at UOG tertiary eye care and training center, North West Ethiopia, 2021.

The prevalence of DR in the worst affected eye was 110 (33.3%), ranging from mild NPDR 30(18.2%) to PDR 3(1.8%). The overall prevalence of DME was 17% of which 11.5% had Non-CSME and 5.5% had CSME in the worst affected eye. (Table-VI)

**Table-VI.** DR and DME grading among diabetic patients at UOG tertiary eye care and training center, North West Ethiopia, 2021 (n=165)

| Variable | Grade of Diabetic retinopathy | Number (%) |
| --- | --- | --- |
| Grade of DR | No DR | 110 (66.70%) |
|  | Mild DR | 30 (18.20%) |
|  | Moderate NPDR | 13 (7.90%) |
|  | Sever NPDR | 9 (5.50%) |
|  | PDR | 3 (1.80%) |
| Grade of DME | No DME | 137 (83.00%) |
|  | DME | 19 (11.50%) |
|  | CSME | 9 (5.50%) |

Seventy one (43%) of patients had visual impairment, out of this 35 (21.2%) had mild visual impairment, 17 (10.3%) had moderate visual impairments and 19(11.5%) were blind.. (Table VII)

**Table-VII.**
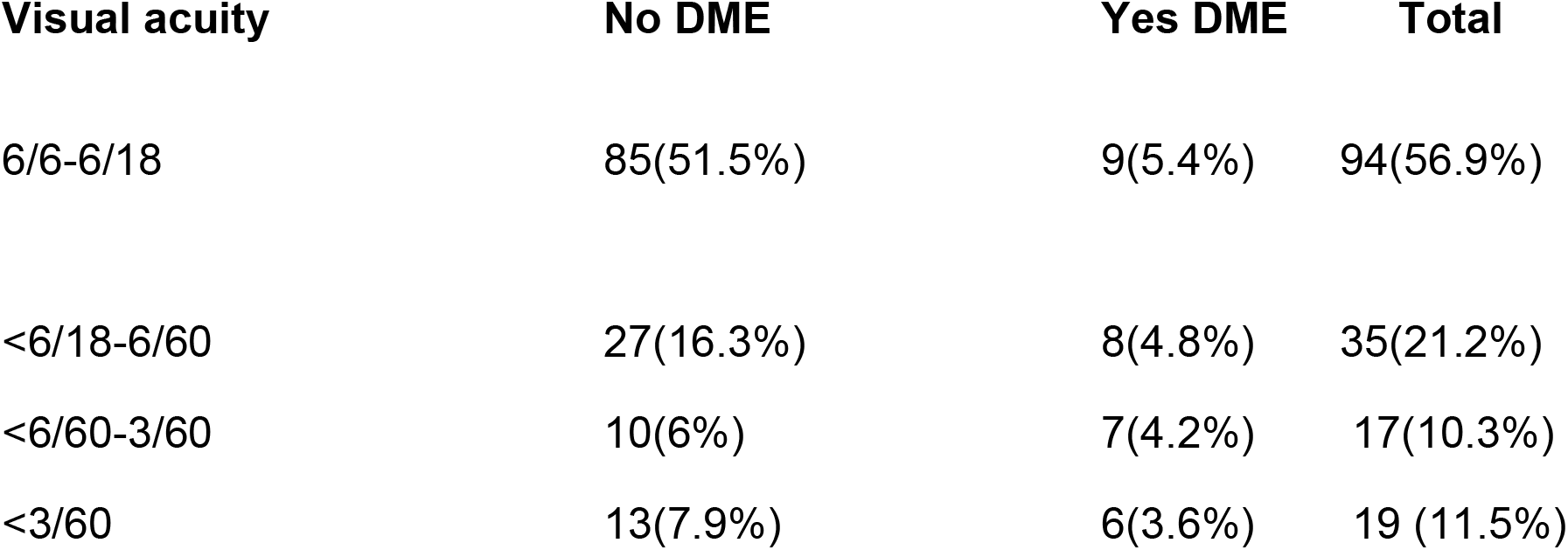
Visual acuity versus the presence or absence of DME among diabetic patients at UOG tertiary eye care and training center, North West Ethiopia, 2021 (n=165)

| Visual acuity | No DME | Yes DME | Total |
| --- | --- | --- | --- |
| 6/6-6/18 | 85(51.5%) | 9(5.4%) | 94(56.9%) |
| <6/18-6/60 | 27(16.3%) | 8(4.8%) | 35(21.2%) |
| <6/60-3/60 | 10(6%) | 7(4.2%) | 17(10.3%) |
| <3/60 | 13(7.9%) | 6(3.6%) | 19 (11.5%) |

A binary logistic regression analysis was done for every independent variable to include into the final multivariable logistic regression model. Then the variables with p-value of less than 0.2 were included into the final model and association of the independent variables with DME.

The bi-variable logistic regression analysis showed that, Residency, Type of DM, Duration of DM, Hypertension, History of cataract surgery, proteinuria and higher Diastolic BP were found to have association (P< 0.2) with outcome (the presence or absence) DME.

In the final multivariable logistic regression analysis patients having proteinuria on urine examination (P<0.01) and those with severe NPDR and PDR (P<0.01) were significantly associated with development of DME.

Accordingly, patients with proteinuria in Urine analysis result were 8.04 times highly likely to develop DME as compared with DM patients with normal Urine analysis result (AOR = 8.04, 95% CI (2.48-26.09).

Similarly DM patients with severe NPDR and PDR were 22.04 times highly likely to have DME as compared with those without DR (AOR = 22.04, 95% CI 2.1-231). (Table VIII)

**Table-VIII.** Multivariate logistic regression of factors associated with DME among diabetic patients at UoG tertiary eye care and training center, North West Ethiopia, 2021 (n=165)

|  |  | Freq. (%) | Freq. (%) | P value | AOR | Lower | Upper |
| --- | --- | --- | --- | --- | --- | --- | --- |
| Residency |  |  |  |  |  |  |  |
|  | Rural | 16(66.7) | 8(33.3) |  |  |  |  |
|  | Urban | 120(85.1) | 21(14.9) | 0.269 | 0.393 | 0.075 | 2.06 |
| Type of DM |  |  |  |  |  |  |  |
|  | Type 1 | 25(73.5) | 9(26.5) |  | 1.321 | 0.242 | 7.216 |
|  | Type 2 | 111(84.7) | 20(15.3) | 0.748 |  |  |  |
| Duration of DM |  |  |  |  |  |  |  |
|  | < 5 years | 62(83.8) | 12(16.2) | 0.084 |  |  |  |
|  | 5-10 years | 41(85.4) | 7(14.6) | 0.651 | 1.49 | 0.265 | 0.651 |
|  | >=10 years | 32(74.4) | 11(25.6) | 0.589 | 1.555 | 0.313 | 7.714 |
| Hypertension |  |  |  |  |  |  |  |
|  | Yes | 51(73.9) | 18(26.1) | 0.66 | 1.392 | 0.319 | 6.073 |
|  | No | 85(88.5) | 11(11.5) |  |  |  |  |
| History of cataract surgery |  |  |  |  |  |  |  |
|  | Yes | 8(57.1) | 6(42.9) | 0.633 | 1.77 | 0.169 | 18.513 |
|  | No | 128(84.4) | 23(15.2) |  |  |  |  |
| Urine analysis |  |  |  |  |  |  |  |
|  | trace and negative | 112(90.3) | 12(9.7) |  |  |  |  |
|  | +1 & above | 11(44.0) | 14(56.0) | 0.01 | 8.04 | 2.481 | 26.095 |
| Diastolic BP |  |  |  |  |  |  |  |
|  | <90mm/Hg | 120(96.8) | 24(92.3) |  |  |  |  |
|  | >=90 mm/Hg | 4(3.2) | 2(7.7) | 0.63 | 0.532 | 0.041 | 6.955 |
| severe NPDR and/or PDR |  |  |  |  |  |  |  |
|  | No | 134(87.6) | 19(12.4%) |  |  |  |  |
|  | Yes | 1(8.3%) | 11(91.7%) | 0.01 | 22.04 | 2.101 | 231.33 |

## Discussion

The overall prevalence of Diabetic Macular Edema and clinically significant macular edema in this study was 17% and 5.5 % respectively. The prevalence of CSME in this study is similar with results of studies from Jima south west Ethiopia and Iran which were 6 %, 5.8%, respectively. (27, 28) The overall prevalence of DME in this study is also in line with results of studies from South Africa, 20.8%, and Turkey, 15.8%. (8,21)

However, the reported prevalence of DME in previous studies in this region of NW Ethiopia, 11%, 6.4% and 5.7%, is lower than our report and this may be because of the differences in the study setting, method of data collection and sample size. (10,11,29) Similar diabetic clinic based studies in Cameroon 8% and in South Africa 12.5%, also reported a lower prevalence of DME than ours. (6,7)

The prevalence of DME in USA and England ranges from 3.8-7.12% which is also lower than this study and this could be due to the differences in sample size, study setting and better medical care and follow up for diabetic patients. (3,5)

A study done in Kenya reported that the prevalence of DME was 33.3% (9) which was much higher than this study and this could be due to the different sampling method used and also included only patients age above 50 years old.

The prevalence of diabetic retinopathy in this study 33.3% is higher than diabetic clinic based previous reports from this region 16 % (10) and 18.9 % (29) however a study done in Jimma South West Ethiopia and another study done in NW Ethiopia reported higher figures than this study, 42.2% and 41.4% respectively. (11,28) This may be due to the differences in data collection technique, sample size and study setting.

The prevalence of DR in our study is higher than reports from other parts of Africa, in Cameroon, 24.3%, in South Africa 24.8% but slightly lower than a Kenyan report 35.9 %. (6,7,9)

The presence of severe NPDR and/or PDR was 22.04 times more likely to have DME than Early or No DR in this study with P value <0.001. A similar finding was reported with a slightly lower figure than ours from Boston USA which was 6.2 times and 7.7 times for severe NPDR and PDR respectively. (13)

A retrospective study of electronic medical records in the UK showed that the presence of any degree of DR was 6.25 times more likely to have DME than absence of DR. (30)

Our study showed that patients with proteinuria in Urine analysis result were 8.04 times more likely to develop DME as compared with DM patients with normal Urine analysis result. Many studies have also shown that the presence of proteinuria had significant association with the development of diabetic macular edema.

In the Wisconsin Epidemiologic Study of Diabetic Retinopathy patients with proteinuria were three times more likely to have DME. (3)

Two retrospective studies done in China and Japan also showed that the presence of microalbuminuria and proteinuria was significantly associated with development of DME. (15,20)

A majority of participants in this study,79.4%, were type-II DM patients and this is similar with previous studies done in Ethiopia, 88.4% in Gondar, 72.8% in Jimma and 60.92% Debre marcos (10,11,28,29)

Longer duration of DM was strongly associated with diabetic macular edema in many studies. (2,3,10,11) However our study did not find association between duration of diabetes with development of DME and this may be because nearly half of our study patients, 44.8%, had duration of diabetes less than five years.

A majority of DM patients in this study were on oral hypoglycemic agents, 49.9%, and 33.9 % of patients were on Insulin alone or combination therapy. Insulin therapy was reported to have significant association with DME in some studies (3,14,31). However, this study didn’t show any significant association.

Poor glycemic control (16,17,19), uncontrolled blood pressure (15,18) and high lipid level (cholesterol level and LDL) (12,15,17) have been associated with the development of DME in some studies but our study did not show any association. The small sample size and our inability to determine HgA1c and lipid level for all patients might have contributed to this discrepancy. These are also the limitations of this study.

The absence of imaging studies like OCT might have also underestimated the prevalence of DME in our patients as the diagnosis of DME was made based on clinical examination only.

## CONCLUSION

The prevalence of Diabetic macular Edema among our patients was very high and this implies the need to establish early screening and proper treatment services to prevent vision loss from DME. The presence of proteinuria was independently associated with the development of diabetic macular edema. Diabetic patients must be taught about the need for regular eye examination to detect and treat DME early.

## Data Availability

All relevant data are within the manuscript and its Supporting Information files.

NA

## REFERENCES

1. Varma R, Bressler NM, Doan QV, Gleeson M, Danese M, Bower JK, et al. Prevalence of and risk factors for diabetic macular edema in the United States. JAMA ophthalmology. 2014;132(11):1334–40.

2. Bhagat N, Grigorian RA, Tutela A, Zarbin MA. Diabetic macular edema: pathogenesis and treatment. Survey of ophthalmology. 2009;54(1):1–32.

3. Klein R, Klein BEK, Moss SE, Davis MD, DeMets DL. The Wisconsin Epidemiologic Study of Diabetic Retinopathy. Ophthalmology. 1984;91(12):1464–74.

4. Xie J, Ikram MK, Cotch MF, Klein B, Varma R, Shaw JE, et al. Association of diabetic macular edema and proliferative diabetic retinopathy with cardiovascular disease: a systematic review and meta-analysis. JAMA ophthalmology. 2017;135(6):586–93.

5. Minassian D, Owens DR, Reidy A. Prevalence of diabetic macular oedema and related health and social care resource use in England. British journal of ophthalmology. 2012;96(3):345–9.

6. Jivraj I, Ng M, Rudnisky CJ, Dimla B, Tambe E, Nathoo N, et al. Prevalence and severity of diabetic retinopathy in Northwest Cameroon as identified by teleophthalmology. TELEMEDICINE and e-HEALTH. 2011;17(4):294–8.

7. Sukha AY, Rubin A. Demographic, medical and visual aspects of Dia-betic Retinopathy (DR) and Diabetic Macular Edema (DME) in South African diabetic patients. African Vision and Eye Health. 2009;68(2):70–81.

8. Webb EM, Rheeder P, Roux P. Screening in primary care for diabetic retinopathy, maculopathy and visual loss in South Africa. Ophthalmologica. 2016;235(3):141–9.

9. Mathenge W, Bastawrous A, Peto T, Leung I, Yorston D, Foster A, et al. Prevalence and correlates of diabetic retinopathy in a population-based survey of older people in Nakuru, Kenya. Ophthalmic epidemiology. 2014;21(3):169–77.

10. Tsegaw A, Alemu S, Dessie A, Patterson CC, Parry EH, Phillips DI, et al. Diabetic Retinopathy in Type 2 Diabetes Mellitus Patients Attending the Diabetic Clinic of the University of Gondar Hospital, Northwest Ethiopia. Journal of Ophthalmology. 2021;2021.

11. Ejigu T, Tsegaw A. Prevalence of Diabetic Retinopathy and Risk Factors Among Diabetic Patients at University of Gondar Tertiary Eye Care and Training Center, North-West Ethiopia. 2021.

12. Yau JW, Rogers SL, Kawasaki R, Lamoureux EL, Kowalski JW, Bek T, et al. Global prevalence and major risk factors of diabetic retinopathy. Diabetes care. 2012;35(3):556–64.

13. Lopes de Faria JM, Jalkh AE, Trempe CL, Mcmeel JW. Diabetic macular edema: risk factors and concomitants. Acta Ophthalmologica Scandinavica. 1999;77(2):170–5.

14. Wang Y, Lin Z, Zhai G, Ding XX, Wen L, Li D, et al. Prevalence of and risk factors for diabetic retinopathy and diabetic macular edema in patients with early and late onset diabetes mellitus. Ophthalmic research. 2020.

15. Tan GS, Cheung N, Simó R, Cheung GC, Wong TY. Diabetic macular oedema. The lancet Diabetes & endocrinology. 2017;5(2):143–55.

16. Bek T. Systemic risk factors contribute differently to the development of proliferative diabetic retinopathy and clinically significant macular oedema. Diabetologia. 2020;63(11):2462–70.

17. Martín-Merino E, Fortuny J, Rivero-Ferrer E, Lind M, Garcia-Rodriguez L. Risk factors for diabetic macular oedema in type 2 diabetes: A case-control study in a United Kingdom primary care setting. Primary care diabetes. 2017;11(3):288–96.

18. Liu E, Craig JE, Burdon K. Diabetic macular oedema: clinical risk factors and emerging genetic influences. Clinical and Experimental Optometry. 2017;100(6):569–76.

19. Eldem B, Ozdek S, Saatci AO, Ozmert E, Ulay E, Nomak G. Clinical Characteristics of Patients with Newly Diagnosed Diabetic Macular Edema in Turkey: A Real-Life Registry Study—TURK-DEM. Journal of ophthalmology. 2017;2017.

20. Zhuang X, Cao D, Yang D, Zeng Y, Yu H, Wang J, et al. Association of diabetic retinopathy and diabetic macular oedema with renal function in southern Chinese patients with type 2 diabetes mellitus: a single-centre observational study. BMJ open. 2019;9(9):e031194.

21. Acan D, Calan M, Er D, Arkan T, Kocak N, Bayraktar F, et al. The prevalence and systemic risk factors of diabetic macular edema: a cross-sectional study from Turkey. BMC ophthalmology. 2018;18(1):1–8.

22. Gabb GM, Mangoni AA, Anderson CS, Cowley D, Dowden JS, Golledge J, et al. Guideline for the diagnosis and management of hypertension in adults - 2016. The Medical journal of Australia. 2016;205(2):85–9.

23. Review executive summary of the clinical guidelines on the identification, evaluation, and treatment of overweight and adult body mass index obesity in adults. Arch Intern Med 1998;158:1855–67.

24. World Health Organization. Consultation on Development of Standards for Characterization of Vision Loss and Visual Functioning. Geneva: World Health Organization; 2003.

25. Liu E, Craig JE, Burdon K. Diabetic macular oedema: clinical risk factors and emerging genetic influences. Clinical and Experimental Optometry. 2017;100(6):569–76.

26. Relhan N, Flynn HW, Jr. The Early Treatment Diabetic Retinopathy Study historical review and relevance to today’s management of diabetic macular edema. Current opinion in ophthalmology. 2017;28(3):205–12.

27. Javadi MA, Katibeh M, Rafati N, Dehghan MH, Zayeri F, Yaseri M, et al. Prevalence of diabetic retinopathy in Tehran province: a population-based study. BMC ophthalmology. 2009;9(1):1–8.

28. Sharew G, Ilako DR, Kimani K, Gelaw Y. Prevalence of diabetic retinopathy in Jimma University Hospital, Southwest Ethiopia. Ethiopian medical journal. 2013;51(2):105–13.

29. Tilahun M, Gobena T, Dereje D, Welde M, Yideg G. Prevalence of Diabetic retinopathy and its associated factors among diabetic patients at Debre Markos referral hospital, Northwest Ethiopia, 2019: hospital-based cross-sectional study. Diabetes, Metabolic Syndrome and Obesity: Targets and Therapy. 2020;13:2179.

30. Colin J Chu 1, Robert L Johnston 2, Charlotte Buscombe 3, Ahmed B Sallam 4, Queresh Mohamed 3, Yit C Yang 5, Risk Factors and Incidence of Macular Edema after Cataract Surgery: A Database Study of 81984 Eyes. Ophthalmology. 2016 Feb;123(2):316–323. doi: 10.1016/j.ophtha.2015.10.001. Epub 2015 Dec 8.

31. Jingi AM, Noubiap JJN, Essouma M, Bigna JJR, Nansseu JRN, Ellong A, et al. Association of insulin treatment versus oral hypoglycaemic agents with diabetic retinopathy and its severity in type 2 diabetes patients in Cameroon, sub-Saharan Africa. Annals of translational medicine. 2016;4(20).

